# MRI signature of brain age underlying post-traumatic stress disorder in World Trade Center responders

**DOI:** 10.1101/2024.10.18.24315761

**Authors:** Azzurra Invernizzi, Francesco La Rosa, Anna Sather, Elza Rechtman, Maryam Jalees, Ismail Nabeel, Alison C. Pellecchia, Stephanie Santiago-Michels, Evelyn J. Bromet, Roberto G. Lucchini, Benjamin J. Luft, Sean A. Clouston, Erin S Beck, Cheuk Y. Tang, Megan K. Horton

**Affiliations:** Department of Environmental Medicine and Public Health, Icahn School of Medicine at Mount Sinai, New York, NY, USA; Department of Neurology, Icahn School of Medicine at Mount Sinai, New York, NY, USA; Department of Artificial Intelligence and Human Health, Icahn School of Medicine at Mount Sinai, New York, NY, USA; World Trade Center Health and Wellness Program, Renaissance School of Medicine at Stony Brook University, Stony Brook, NY, USA; Department of Psychiatry, Renaissance School of Medicine at Stony Brook University, Stony Brook, NY, USA; Department of Environmental Health Sciences, Robert Stempel School of Public Health, Florida International University, Miami, Florida, USA; Department of Medicine, Renaissance School of Medicine at Stony Brook University, Stony Brook, NY, USA; Program in Public Health and Department of Family, Population, and Preventive Medicine, Renaissance School of Medicine at Stony Brook University, Stony Brook, NY, USA; Department of Radiology and Psychiatry, Icahn School of Medicine at Mount Sinai, New York, NY, USA; Department of Public Health and General Preventive Medicine, Internal Medicine, Mount Sinai, New York, NY, USA

**Author notes:** Correspondence: A. Invernizzi at Horton Laboratory. Department of Environmental Medicine and Public Health, Icahn School of Medicine at Mount Sinai, One Gustave Levy Place, Box 1057, New York, NY, 10029, USA. these authors equally contributed to this work (Azzurra Invernizzi, Francesco La Rosa).

**Keywords:** structural MRI, brain age, convolutional neural network, World Trade Center responders, post- traumatic stress disorder, exposure

## Abstract

The men and women involved in rescue and recovery operations at the 9/11 World Trade Center (WTC) site have a greater prevalence (23%) of persistent, clinically significant post-traumatic stress disorder (PTSD). Recent structural and functional magnetic resonance imaging (MRI) studies demonstrate significant neural differences between WTC responders with and without PTSD. Here, we used brain age, a novel MRI-based data-driven biomarker optimized to detect accelerated structural aging, and examined the impact of PTSD on this process. Using BrainAgeNeXt, a novel convolutional neural network trained and validated on 11,574 magnetic resonance imaging (MRI) T1- weighted scans, we predicted brain age in WTC responders with PTSD (WTC-PTSD, *n* = 47) and age/sex matched responders without PTSD (non-PTSD, *n* = 52). Predicted Age Difference (PAD) was then calculated for each WTC responder by subtracting chronological age from brain age. A positive PAD indicates that the responder’s brain is aging faster than expected for their chronological age. We found that PAD is significantly greater with WTC-PTSD compared to non-PTSD responders (*p* < 0.001). Further, we found that WTC exposure duration (months working on site) moderates the association between PTSD and PAD (p=0.0050). Our results suggested that brain age is a valid biomarker to compare aging trajectories in responders with and without PTSD. In particular, PTSD may be a substantial risk factor for accelerated neurodegeneration in this vulnerable and aging population.

## Introduction

The individuals who participated in the rescue and recovery efforts at the World Trade Center (WTC) site following 9/11 attacks have a notably higher prevalence (23%) of chronic, clinically significant post-traumatic stress disorder (PTSD)^1,2^. PTSD is a neuropsychiatric condition triggered by the experience of traumatic events, characterized by hypervigilance and hyperarousal, intrusive and recurrent flashbacks or dreams, and psychological and physiological distress in response to trauma-resembling cues^3^. Among WTC responders and more generally, in the PTSD patient population, PTSD has been linked to several epigenomic, immune and inflammatory biomarkers indicative of accelerated cellular aging, compared to chronological aging^30–40^. Individuals with PTSD have been identified as being at increased risk of age-related medical conditions, such as premature cognitive decline^6,7^. Individuals diagnosed with PTSD early in life (<40 years old) are twice as likely to develop Alzheimer’s later in life^6,7^ with neurodegenerative pathological signatures including hippocampal volume loss^8–11^, cortical thinning^12–14^, changes in white matter connectivity, and amyloidosis^15^. These brain changes indicating pathological aging culminate to increased morbidity and mortality^16^, posing a significant health threat to this already vulnerable population. These brain changes are especially notable in the population of WTC responders, who face an exacerbated prevalence of premature cognitive impairment and its neuropathological underpinnings in mid-life – decades earlier than when age-expected neurocognitive decline typically is observed^17–19^.

Recent neuroimaging studies of WTC responders, including those from our group, leverage a diverse array of MRI modalities to investigate structural brain changes^20–22^ and functional neural-mechanisms^22–27^, such as reduced cortical complexity^22^, that may mediate cognitive differences observed between WTC responders with PTSD (WTC-PTSD) and those without PTSD (non-PTSD). Several studies have shown that responders with greater exposure to the disaster site and more severe PTSD symptoms are at even higher risk for cognitive impairment^17,27,28^ and associated neuropathology^27,29^, such as cortical atrophy. All together these findings further increase the health burden on responders with comorbid PTSD and cognitive impairment. Yet, the identification of a whole-brain aging biomarker applicable to the population of WTC responders has yet to be established.

The pressing need to disentangle PTSD and its impact on the trajectory of healthy brain aging necessitates the identification of prodromal biomarkers sensitive to accelerated aging and related neurodegeneration. This prodromal biomarker will help to inform diagnosis, prognosis, and treatment intervention. “Brain age” is an estimate of the biological age of the brain based on magnetic resonance imaging (MRI) scans. MRI-based brain age has proven to be a valuable marker for identifying accelerated aging, particularly by detecting subtle brain atrophy and early neurodegenerative changes that may not be found in a standard clinical assessment^41^. Here, we apply a state-of-the-art whole-brain MRI-based brain age estimation method sensitive to accelerated brain aging across the entire human lifespan. Among WTC responders with and without PTSD, we calculate and compare responders’ “brain age” to chronological age to derive “predicted age difference (PAD)”^42,43^, a prodromal biomarker of individual’s brain aging with reference to normative brain aging curvatures. Using “PAD”, we compare neurological aging signatures between responders with and without PTSD, identify brain regions associated with PAD, and gain insight into the neural mechanisms driving accelerated brain aging in PTSD. Finally, we examine how the association between PAD and PTSD is moderated by the duration of time responders were exposed to rescue and recovery sites. This study expands our knowledge of the neurological basis linking PTSD and brain aging, highlighting the impact of exposures at 9/11 on the human brain in WTC-responders.

## Methods and Materials

### Participants

Ninety-nine World Trade Center (WTC) responders were recruited from a single clinic associated with the CDC/NIOSH WTC Health Program. This program monitors workers and volunteers from three WTC-exposed cohorts in New York City, all of whom were involved in rescue, response, recovery, cleanup, and related activities following the 9/11 attacks^44,45^. Complete details of the study can be found in previous publications^17,27,45,46^. All participants were 44 to 65 years of age, fluent in English, and satisfied eligibility criteria for MRI scanning (i.e., metal implants or shrapnel, claustrophobia, no prior history of traumatic brain injury, body mass index (BMI) ≤ 40). Global cognitive status was assessed using the Montreal Cognitive Assessment (MoCA)^17^. Diagnostic and assessment of PTSD was determined from the Structured Clinical Interview for the DSM-IV (SCID-IV), described in detail below. Upon enrollment, case (WTC-PTSD) and control (non-PTSD) groups were matched based on age within 5 years, sex, race/ethnicity, occupation (i.e., police) and education at the time of 9/11. Complete demographic information including age, sex, MoCA scores, comorbidities, medications, and WTC exposure are reported in Table 1.

**Table 1.** - Sociodemographic and clinical characteristics of WTC responders selected into the current study (N=99).

| Characteristics | All WTC responders (n=99) | PTSD- (n=52) | PTSD+ (n=47) | p |
| --- | --- | --- | --- | --- |
| **Age (n)** |  |  |  | 0.33 |
| mean ± sd | 55.87 ± 5.22 | 56.36 ± 5.34 | 55.34 ± 5.07 | |
| **Sex (n, %)** |  |  |  |  |
| Male | 79 (78%) | 40 (51%) | 39 (49%) |  |
| Female | 20 (22%) | 12 (60%) | 8 (40%) |  |
| **MoCA (n)** |  |  |  | 0.971 |
| mean, [range] | 23.23 [12,30] | 23.25 [15,30] | 23.21, [12,30] |  |
| **Comorbidities (n, %)** |  |  |  |  |
| Major Depressive Disorder |  |  |  | <0.001* |
| No | 81 (82%) | 52(100%) | 29 (60%) |  |
| Yes | 18 (18%) | 0 (0%) | 18 (40%) |  |
| Cognitive Impairment |  |  |  | 0.989 |
| No | 51 (51.5%) | 27 (53%) | 23 (49%) |  |
| Yes | 48 (48.5%) | 25 (47%) | 24 (51%) |  |
| **WTC Exposure (mean ± sd)** |  |  |  | 0.771 |
| Total months on site | 2.73 ± 5.1 | 2.58 ± 5.2 | 2.88 ± 4.96 | |
| **DSM-IV SCID Trauma Screen (mean ± sd)** |  |  |  |  |
| Re-experiencing | 17.48 ± 6.73 | 12.17 ± 3.12 | 23.4 ± 4.29 | <0.001* |
| Avoidance | 24.01 ± 9.54 | 16.27 ± 3.23 | 32.71 ± 6.12 | <0.001* |
| Hyperarousal | 17.76 ± 6.73 | 12.13 ± 2.92 | 23.97 ± 3.44 | <0.001* |
| Negative thoughts | 12.23 ± 4.58 | 9.04 ± 1.66 | 15.76 ± 4.18 | <0.001* |

**Note:** Mean, standard deviation (sd), range (minimum and maximum values), and percentage (%) are reported. P-values quantify differences between WTC-PTSD and non-PTSD were derived using Student *t*-tests for continuous variables and tests for categorical variables. \* $p<0.005$ . MoCA, Montreal Cognitive Assessment; Cognitive assessment defined as MoCA scores <20/30 point.

Study procedures that follow the Declaration of Helsinki, were approved by the Institutional Review Boards at both Stony Brook University and the Icahn School of Medicine at Mount Sinai. All participants signed informed consents upon explanation of study procedures prior to enrollment.

### Neurological assessments

Clinical assessment of global cognitive functioning was done administering the Montreal Cognitive Assessment (MoCA)^17^ within three months prior to scanning. Cognitive status was reconfirmed at the time of MRI scanning. PTSD diagnosis was assessed using the Structured Clinical Interview for the DSM-IV (SCID-IV)^49^, a semi-structured interview administered by trained clinical interviewers. The PTSD module used WTC exposures as the index trauma. Eligibility criterion for PTSD status was presence (PTSD)/absence (non-PTSD) of current PTSD diagnosis. Symptom subdomains were measured using subscales calculated using reported symptom severity in the SCID for the following symptom domains: re-experiencing, avoidance, hyperarousal, and negative thoughts symptoms (scores ranging from [10-30], [14- 42], [10-30], [8-24], respectively). Major depressive disorder was assessed using the SCID-IV and the presence or absence of current (i.e., active in the past month) major depressive disorder (MDD) diagnosis was determined. MDD was not an exclusion criterion.

### WTC exposure duration

Upon enrolment into the WTC General Responders Cohort^45^, all responders completed a questionnaire about their experience during rescue and recovery efforts. WTC exposure was calculated from self-reported responses to items regarding the time spent (expressed in months and collected at enrollment in the parent study) working on the WTC sites^18,50^. This exposure variable was not available for 10 participants in our study, therefore analyses including this variable were done using a sample of 89 WTC responders. There is no significant difference between the participants removed (n=10) and the participants included in the analysis (n=89) in any demographic characteristics. There was also no group difference in PTSD status (Table 1).

### MRI data acquisition and preprocessing

Magnetic resonance imaging (MRI) data acquisition was performed on a 3-Tesla SIEMENS mMR Biograph scanner using a 20-channel head and neck coil. For each WTC responder, a high-resolution 3D T1-weighted structural brain scan was acquired using a MPRAGE sequence (TR=1900ms, TE=2.49ms, TI=900ms, flip angle-9, acquisition matrix=256x256 and 224 slices with final voxel size=0.89x0.89x0.89 mm). All T1w images underwent skull-stripping using SynthStrip^51^, N4 bias field correction with default parameters, and linear registration with 6 degrees of freedom to the MNI152 1mm isotropic space with ANTs^52^. This minimal preprocessing ensures that all anatomical images are in the same space, enabling consistent analysis across subjects.

### Brain age prediction: BrainAgeNeXt

To predict brain age, we applied the BrainAgeNeXt model^53^, a novel convolutional neural network (CNN) designed for accurate brain age estimation from T1-weighted MRI scans. BrainAgeNeXt is based on the MedNeXt architecture, a transformer-inspired CNN developed specifically for 3D medical images^54^. The original MedNeXt segmentation network has been adapted by using only the encoder path for the brain age regression task. The model automatically extracts relevant features from the entire 3D volume of T1-weighted MRI scans, offering a significant advantage over older machine learning methods that rely on pre-extracted brain volumetric features^55^. BrainAgeNeXt^53^ was trained on a large dataset of 11,574 1.5T and 3T MRI scans from healthy individuals, aged 5 to 95 years, who were imaged at 75 sites. The method consists of five individual models, all trained in the same way with randomly initialized weights, and the final output is obtained by combining their predictions using a mean ensemble approach. On a separate testing set of 1,523 MRI scans, BrainAgeNeXt achieved a mean age error of 2.78 ± 3.64 years, outperforming state-of-the-art methods^56,57^. In our study, each subject’s T1-weighted scan was processed through BrainAgeNeXt to obtain the predicted brain age. We then calculated the difference between brain age and chronological age to determine the Predicted Age Difference (PAD) as it serves as a quantitative marker of brain aging.

## Data availability

De-identified data will be made available upon reasonable request to the corresponding author; raw image files can be accessed upon completion of a data use agreement.

## Statistical Analysis

All analyses were conducted in the R statistical software package. To examine differences in clinical and demographic characteristic across groups (WTC-PTSD and non-PTSD), pairwise Student *t*-test with Welch’s correction for continuous variables and _2_ tests for categorical variables were used. To assess model performance, we calculated Pearson correlation coefficient (R) and the explained variance of the model (VE) between chronological age and predicted brain age. Brain PAD (predicted brain age - chronological age) was calculated for each subject, using BrainAgeNeXt, and was used as the primary outcome measure. To quantify differences in predicted age and PAD between WTC responders with and without PTSD, pairwise Student *t*-tests were used. To better understand BrainAgeNeXt predictions, we investigated monotonic correlations between brain volumes as obtained using SynthSeg^58^ based on FreeSurfer^56^, a segmentation approach based on CNN trained with synthetic data, and PAD were evaluated using Spearman’s rank correlation in all 99 WTC-responders and then, stratified by responders with PTSD (WTC-PTSD) and without PTSD (non-PTSD; Figure 2). We combined brain volumes from the left and right hemispheres, and each volume was normalized by the total intracranial volume. Statistical significance was determined using a *p*-value threshold of 0.05, adjusted for multiple comparisons using the false discovery rate (FDR) correction. Finally, to test our hypothesis that WTC exposure duration (months on site) moderated the association between PTSD and PAD, general linear model (GLM) regressions were computed using current PTSD diagnosis and cumulative WTC exposure duration expressed in months as predictors and PAD as the outcome.

## Results

### Demographic and clinical characteristics

Table 1 reports the clinical and demographic characteristics for the 99 WTC responders included in this study stratified by responders with PTSD (WTC-PTSD) and without PTSD (non-PTSD). Responders were in their mid-fifties at the time of the imaging data acquisition (55.8 ± 5.2 years) and the majority were male (78%). By design, groups were matched on age at the time of the neuroimaging scanning, sex, race/ethnicity, and educational attainment. Current major depression diagnosis (MDD) and PTSD symptoms scales (DSM-IV SCID trauma screen) differed significantly differ between groups: WTC-PTSD has a significantly higher prevalence of MDD compared to non-PTSD (40% vs 18%); higher reported PTSD scales in WTC-PTSD (Table 1). No significant difference in WTC-exposure duration was found between WTC responders with/without PTSD (average month on site for WTC-PTSD= 2.88 and non-PTSD=2.58, p=0.771).

### Prediction of brain age in WTC responders

Predicted brain age from our BrainAgeNeXt model and chronological age were significantly correlated (*R*=0.77 and VE=59.31%). By design, sample subgroups were matched on chronological age at the time of scan thus there were no differences in chronological age between WTC-PTSD and non-PTSD (*p* = 0.33; Table 2 and Figure 1, panel A). No significant difference was observed in predicted brain age between WTC-PTSD and non-PTSD (*p* = 0.099; Table 2 and Figure 1, panel B). PAD differed significantly between WTC-PTSD and non-PTSD responders where WTC-PTSD shown a higher mean PAD of 3.07 years compared to non-PTSD responders, who had a mean PAD of -0.43 years, *p* < 0.001 (Table 2 and Figure 1, panel C).

**Figure 1.**
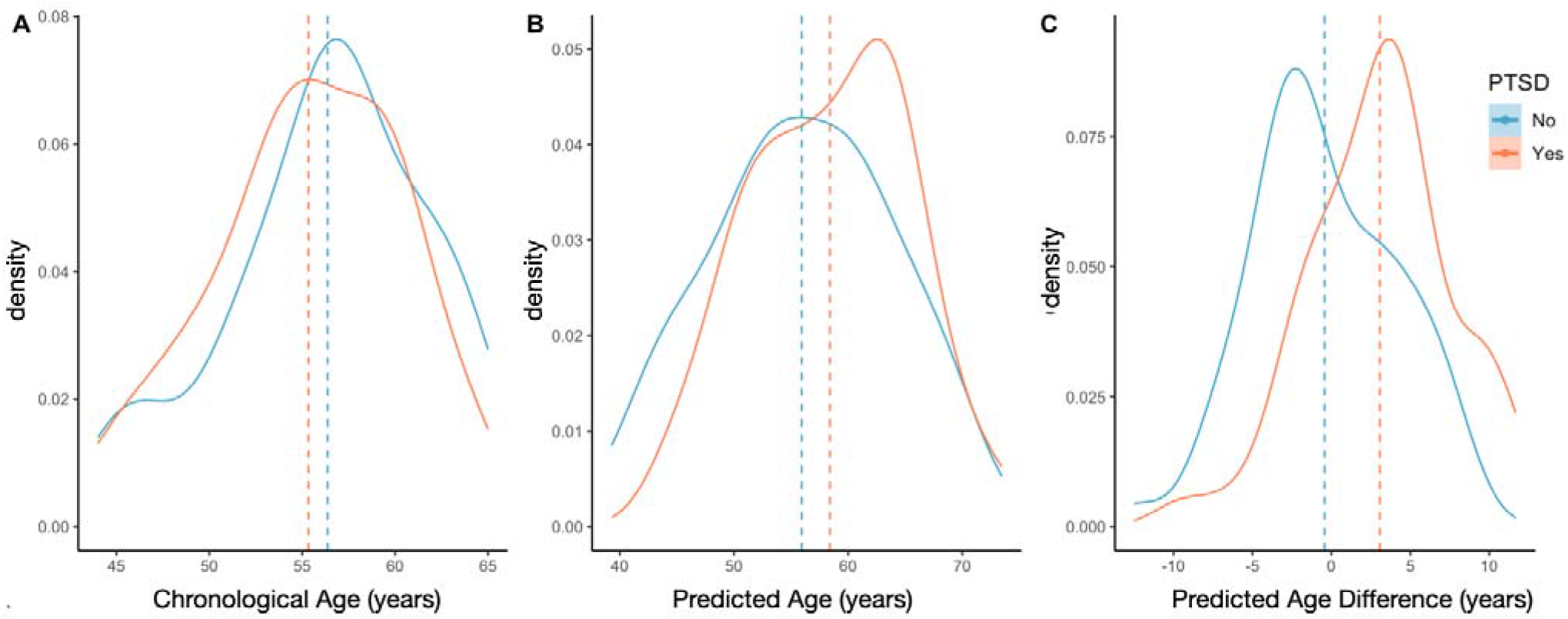
- Predicted Age Difference (PAD) is greater in WTC-PTSD responders. Density plots showing the distribution of chronological age (Panel A), predicted brain age (Panel B) and Predicted Age Difference (PAD; Panel C) stratified by WTC-PTSD (orange line) and non-PTSD (blue line). Only PAD differed significantly between WTC-PTSD and non-PTSD groups.

**Figure 2.**
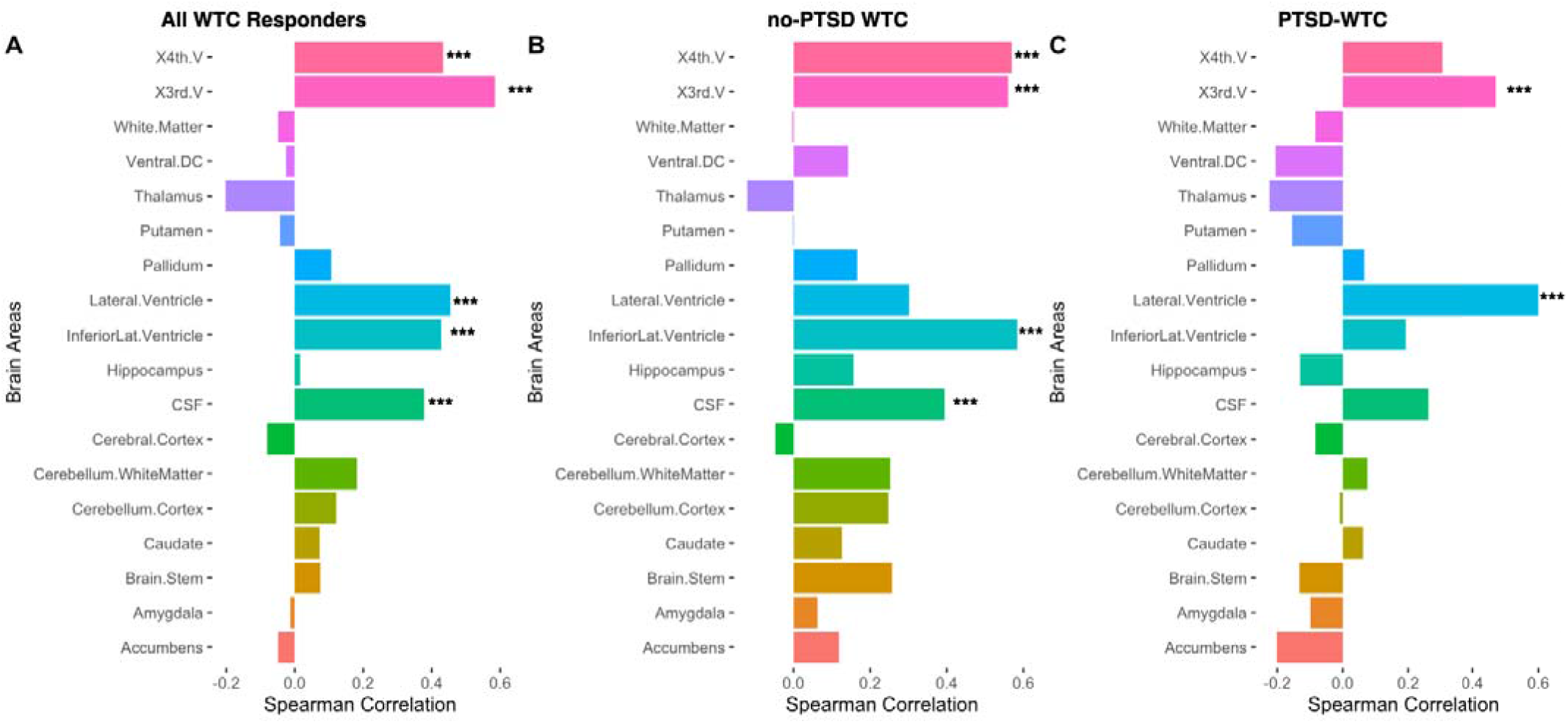
PAD strongly correlates with specific brain areas. Correlations between brain volumes (normalized for intracranial brain volume) and PAD for all 99 WTC responders (Panel A), non-PTSD (Panel B) and WTC-PTSD (Panel C). Brain parcellation was performed with SynthSeg (FreeSurfer)^58^. Significance level: * *p* < 0.05, ** *p* < 0.01, *** *p* < 0.001.

**Table 2.** - Statistical differences in chronological age, predicted brain age and predicted age differences (PAD) between WTC-PTSD and no-PTSD groups.

| Characteristics (years) | All WTC responders (n=99) | PTSD- (n=52) | PTSD+ (n=47) | p |
| --- | --- | --- | --- | --- |
| **Age** |  |  |  | 0.330 |
| mean ± sd | 55.87 ± 5.22 | 56.36 ± 5.34 | 55.34 ± 5.07 | |
| **Predicted Age** |  |  |  | 0.099 |
| mean ± sd | 57.12 ± 7.51 | 55.93 ± 7.92 | 58.41 ± 6.88 | |
| **PAD** |  |  |  | <0.001* |
| mean ± sd | 1.23 ± 4.82 | -0.43 ± 4.54 | 3.07 ± 4.48 | |

**Note:** P-values quantify differences between WTC-PTSD and non-PTSD were derived using Student *t*-tests.

### Brain volumes and PAD

Across All WTC-responders, PAD was most strongly correlated with the following brain areas: fourth ventricle (*r* = 0.424, *p* < 0.001), third ventricle (*r* = 0.621, *p* < 0.001), lateral ventricle (*r* = 0.461, *p* < 0.001), inferior lateral ventricle (*r* = 0.452, *p* < 0.001) and cerebrospinal fluid (*r* = 0.424, *p* < 0.001). For non-PTSD responders, PAD was significantly correlated with the fourth ventricle (*r* = 0.615, *p* < 0.001), third ventricle (*r* = 0.611, *p* < 0.001), inferior lateral ventricle (*r* = 0.519, *p* < 0.001) and cerebrospinal fluid (*r* = 0.51, *p* < 0.001) For WTC-PTSD responders, PAD correlated significantly with the third ventricle (*r* = 0.591, *p* < 0.001) and lateral ventricle (*r* = 0.581, *p* < 0.001). Correlations between brain volumes and predicted brain age are reported in supplementary materials (Figure S1). Brain volumes that were significantly correlated with PAD and/or predicted brain age metrics were not significantly different between responders with and without PTSD (Table S1).

### Impact of WTC exposure on PAD

Results from generalized linear modeling suggest that WTC exposure duration (months on site) moderates the association between PTSD and PAD (p-value for interaction = 0.050, 95% CI [-0.0, 1.12]; Figure 3). In WTC-PTSD responders, prolonged WTC exposure is associated with increased PAD. For completeness, models that used chronological and predicted age as outcome were reported in supplementary materials (Figure S2).

**Figure 3.**
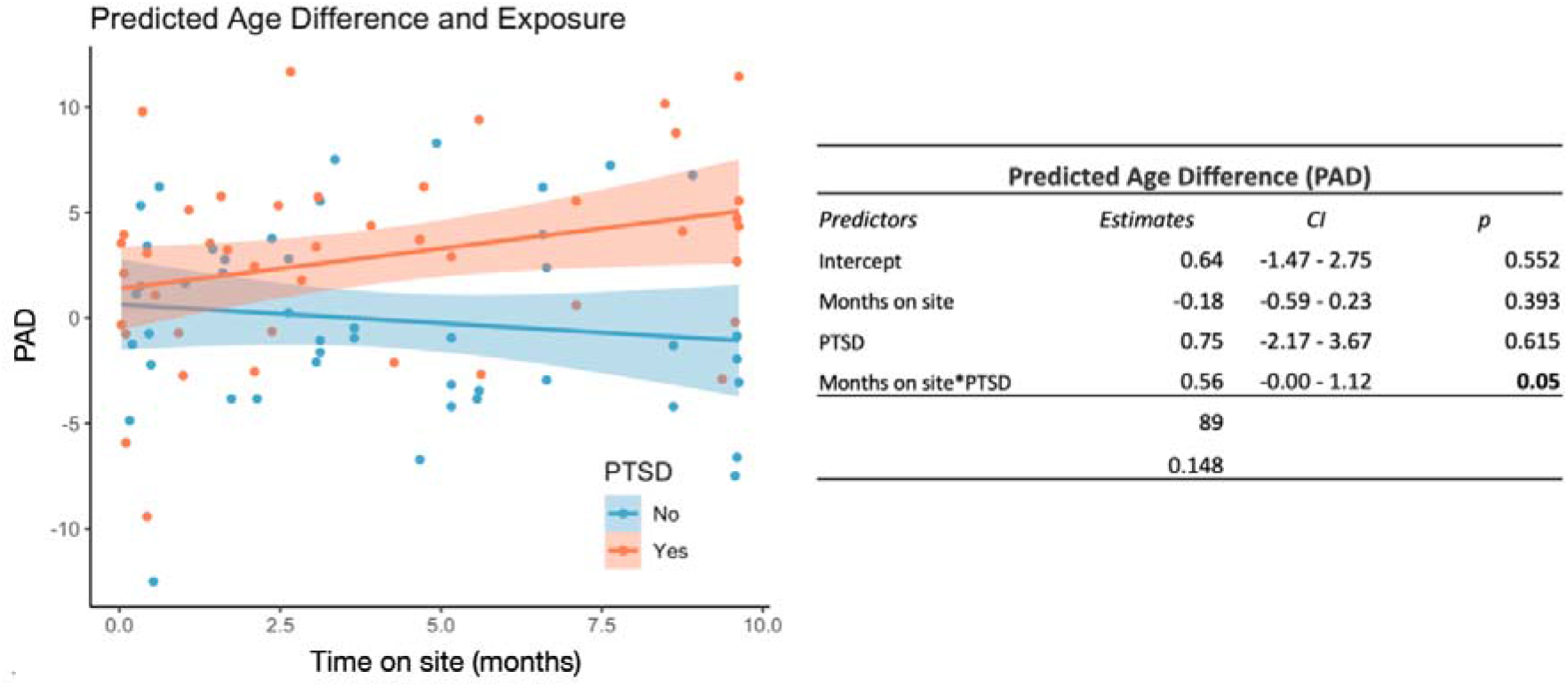
- WTC exposure duration moderates association between PTSD and PAD. This graph plots the relationship (interaction) between WTC exposure duration in months (x-axis) and predicted age difference (PAD) (y-axis) stratified by WTC-PTSD (orange dots) and non-PTSD (blue dots). WTC exposure duration (months on site) moderates the association between PTSD status and PAD (p-value for interaction = 0.050, 95% CI [-0.0, 1.12].

## Discussion

MRI-based brain age models are key to understanding healthy aging trajectories, identifying deviations from these patterns, and identifying prodromal biomarkers that may indicate early-stage neurodegeneration^56^. These models provide valuable insights into the brain’s structural changes, helping to link early signs of neurodegenerative diseases with long-term clinical outcomes. In this study, we investigated the relationship between traumatic exposures, PTSD, and brain age among WTC responders. Using a novel convolutional neural network (CNN), we compared chronological age to predicted-brain age, estimating the PAD, an imaging biomarker derived from anatomical MRI that captures structural brain variations with high precision for each WTC responder. We found a significant increase in PAD among responders with PTSD compared to those without (p < 0.001), providing evidence that those with PTSD may endure premature aging compared to those without PTSD. Additionally, the duration of WTC exposure (months on site working in rescue and recovery efforts) moderated the association between PTSD and PAD (p = 0.0050). These findings provide a neurological basis for linking PTSD and brain aging, highlighting brain areas that may be associated with greater PAD among responders with PTSD. This suggests that PTSD is a substantial risk factor for neurodegeneration, accelerating brain aging – a process moderated by the degree of exposure to the rescue and recovery site. Our findings also provide support for the use of “brain age” and “PAD” as promising diagnostic and prognostic biomarkers for neurodegeneration in the broader vulnerable population of aging individuals diagnosed with PTSD.

Brain age significantly differs between WTC responders with and without PTSD, with those experiencing WTC-related PTSD showing accelerated brain aging, as reflected by higher Predicted Age Difference (PAD) estimates - the gap between brain predicted and chronological age (Table 2, Figure 1). Previous studies have already linked PTSD among WTC responders to anatomical^22,25^ and functional neural changes^59^, as well as a range of aging-related conditions manifesting clinically as memory loss^18^, increased risk of cognitive impairment^60–62^ and physical functional limitations^50^. Additionally, gene expression^63^ and DNA methylation^40^ metrics of epigenetic and transcriptomic aging indicate that WTC responders with PTSD are aging at significantly accelerated rate compared to those without PTSD^34^. These structural, functional, cognitive and cellular changes provide a clear link between PTSD and the accelerated aging consistent with our WTC-PTSD responder sample. Taken together, these findings highlight the profound impact of PTSD on long-term brain health, underscoring that PTSD is not only a chronic psychological condition but also a significant contributor to accelerated biological aging^37,64^, with its underlying neural mechanisms playing a key role in this process.

As our population ages, there is an increasing need to understand where and how the brain changes over time and the clinical implications of these changes^65^. A notable strength of our study was using a convolutional neural network (CNN) to derive brain age^53^. One of the key advantages of BrainAgeNeXt is its ability to learn directly from 3D MRI data without relying on pre-extracted volumetric features. While this approach mitigates the issues associated with brain parcellation variability and facilitates the model’s applicability to real-world clinical settings, a limitation is understanding the neural drivers of prediction. To overcome this, we explored which brain area volumes were most correlated with PAD. Among WTC responders, we found that PAD was correlated with volumes of several key brain structures (Figure 2). Specifically, the third, fourth, lateral, and inferior lateral ventricles, as well as cerebrospinal fluid (CSF). Notably, among responders with PTSD (WTC-PTSD), the third and lateral ventricles showed the strongest positive correlations with PAD. In contrast, for responders without PTSD (no-PTSD), the third, fourth and inferior lateral ventricles and CSF were most strongly correlated with PAD. These findings align with well-established aging patterns, where increased CSF volume reflects brain atrophy, including cortical shrinkage and ventricular enlargement^66^. As individuals age, ventriculomegaly^67^, a process involving the enlargement of brain ventricles, occurs as the surrounding gray matter tissues experience a reduction in volume^68^. This volumetric increase in CSF is typical of normal aging, indicating brain atrophy,^69,70^ but is often more pronounced in neurodegenerative conditions^71–73^. Our findings in all WTC responders are consistent with these established neuroanatomical changes in aging populations, where increased PAD corresponds to more pronounced age-related brain atrophy. Finally, as interest grows in identifying methods to monitor increased risk of neurodegeneration conditions, our findings establish that brain age is a valuable biomarker for detecting neurodegenerative processes in PTSD. Our findings underscore the need for continued research in this area. Future studies should explore whether therapeutic interventions in WTC responders can slow the rate of accelerated aging in those with PTSD. Additionally, there is a critical need to investigate the relationship between the influence of trauma exposure and PTSD severity on accelerated brain aging. Understanding these connections could pave the way for targeted interventions to promote healthier brain aging in populations affected by PTSD.

We investigated the correlation between the predicted brain age and brain volumes. Interestingly, the thalamus emerged as the only non-ventricular brain region negatively associated with increased predicted age in WTC-PTSD responders but not in those without PTSD (Figure S1). Notably, none of these brain volumes showed statistically significant differences between responders with and without PTSD (Table S1). The thalamus plays a key role in sensory, emotional, and fear regulation, acting as a relay between the amygdala and prefrontal cortex. In animal models, the thalamus showed vulnerability to the cumulative effects of both psychological and environmental stress^74^. Neuronal cell death was observed in brain areas, including the thalamus, only among rats with combined exposure to stress and low doses of environmental neurotoxins^74^. Reduced thalamic volume has been observed in structural MRI studies of PTSD, consistent with our finding that smaller thalamic volumes are linked to greater predicted age in responders with PTSD^75,76^. Among the WTC cohort, previous studies identified reduced white matter integrity in the thalamic radiation, a connection between the thalamus to the cerebral cortex^77^. Further studies are necessary to explore the specific role of the thalamus and other brain structures most vulnerable to this decline, which may become key targets for clinical therapies aimed at mitigating and slowing the accelerated neurodegenerative processes.

Among the more than 35,000 responders enrolled in the ongoing WTC-HP, 23% of them continue to experience chronic WTC-related PTSD^1,16,78^. In this study of responders selected on PTSD case status (WTC-PTSD vs WTC non-PTSD), the duration of WTC exposure (i.e., number of months spent at the WTC site in rescue and recovery efforts) did not differ between responders with PTSD and those without PTSD (Table 1). While all responders experience some degree of traumatic exposure, not all responders develop PTSD. Among responders with PTSD, greater PAD is associated with prolonged WTC exposure during search and rescue efforts at and for months after 9/11 (Figure 3), suggesting that prolonged exposure may exacerbate the already adverse effects of PTSD on brain aging in this vulnerable population. During these months on the pile, WTC responders experienced psychologically traumatic stressors and inhaled dust and smoke containing many pollutants (i.e., particulate matter, lead, polycyclic aromatic hydrocarbons (PAHs), polychlorinated biphenyls (PCBs), and dioxins). Circulating particulates and/or toxicants may reach the brain through the olfactory system, disrupting blood-brain barrier function, and inducing neuroinflammation at the global-network^18,50^. This unique combined exposure to traumatic stressors and environmental toxicants may play a role in the anatomical and functional changes observed in our population^79–82^. Our results are consistent with previous studies within the WTC cohort that found associations between longer-duration WTC exposure and greater structural changes such as reduced cortical volumetric and decreased cortical complexity^22,25^. Synthesizing our results and prior literature, there may be a synergistic interaction between PTSD and prolonged exposure to psychological and environmental stressors at play, amplifying a responders susceptibility to neurodegeneration.

Our findings demonstrate the feasibility of using a whole-brain MRI-based "brain age" biomarker to identify and quantify deviations from healthy aging trajectories, assessing an increased risk of neurodegeneration. "Brain age" is a well-validated measure in healthy individuals and has been extensively studied in populations with neurodegenerative diseases^83–85^ and neuropsychiatric disorders^53,86–88^. However, the application of "brain age" to PTSD is largely underexplored. While recent studies suggest a potential link between PTSD and accelerated brain aging as measured by PAD^89,90^, these findings have been inconsistent and suffer from methodological biases. One study found preliminary evidence that PTSD may be associated with accelerated brain aging^90^, but it had several limitations: a younger participant sample, imbalanced group sizes, and a control group that lacked trauma exposure. Another study reported that young males with PTSD had a higher PAD compared to both young and old male controls^89^. However, older males with PTSD exhibited a lower PAD, contradicting growing evidence that PTSD likely contributes to accelerated brain aging^89^. Like the previous study^90^, this one was also limited by the young and broad age range of participants, with an average age of 35.6 years, a stage at which significant neurodegenerative changes are generally not prominent, making it difficult to accurately capture PAD^89^. Despite these intriguing preliminary results, the underlying mechanisms by which psychological trauma, such as PTSD, might disrupt healthy brain aging—particularly at a neurological level—remain unclear. Our study sample and design presented a unique opportunity to examine brain aging in regards to a homogenous traumatic experience, the 9/11 World Trade Center tragedy and its aftermath. Having an age-matched control group without PTSD who experienced the same trauma as their counterparts who with PTSD, allows us to isolate the effects of the psychological and physiological sequelae of PTSD rather than the less specific effects of trauma-exposure or environmental toxic exposures. This minimizes potential confounding external variables between groups, providing more support for a causal inference and overall validity of our results.

Limitations of our study design include the small sample size and lack of an external control group (non-WTC). A larger sample size might improve statistical power. While a strong effort was made to increase the recruitment of underrepresented populations including women and people of color to the point of doubling the numbers of both in this sample compared to the responder population enrolled in our program^27,50,60^, our sample could nevertheless benefit from improved diversity in order to facilitate subgroup analyzes that are out of reach of this study. The age range of WTC-responders included in this study (mean age of 55 years, ranging from 44 to 65) might be considered a limitation to the generalizability of our results as other PTSD studies typically focus on younger-aged groups rather than late adulthood. However, less is known about the effect of PTSD on the brain in individuals in this later stage of life and there is a need for more studies in this understudied age category. The exposure questionnaire, from which we gather self-reported experience during WTC rescue and recovery efforts, was often first administered years after 9/11 experiences were completed and may therefore be subject to recall bias. Finally, we lack accurate assessments of life trauma and/or PTSD status and MRI scans in WTC responders prior to 9/11, and we lack a comparison group of responders with subsyndromal, mild, heterogeneous, or remitted PTSD. While these limitations do reduce the generalizability of these findings in the general population, individuals exposed to traumatic circumstances are always different in critical ways from the general population. In our study, the relationship between PTSD and brain age was assessed at a single time point, cross sectionally. Thus, causal inferences cannot be drawn for the association and directionality of the PTSD and possible neurodegeneration interaction. Future studies would benefit from examining PTSD and brain aging longitudinally, providing insight into the temporality of this association.

To conclude, this is the first study to investigate the neurological basis linking PTSD and brain aging, highlighting the impact of traumatic stressors in WTC responders. We successfully derived Predicted Age Differences (PAD) from MR imaging using a novel convolutional neural network to compute brain age in a PTSD population. Our results suggest that responders who developed WTC-related PTSD present significantly accelerated brain aging compared to non-PTSD responders. PAD is correlated with volumes of specific brain areas previously shown to be associated with aging and PTSD. Finally, brain age is associated with WTC-exposure. By offering a reliable biomarker to assess aging trajectories, this study expands our understanding of the neurobiological impact of PTSD in WTC responders and highlights the broader potential of “brain age” to inform targeted individual and public health interventions aimed at promoting healthy brain aging across the lifespan.

## Acknowledgements

The authors would like to acknowledge support from the Centers for Disease Control and Prevention for supporting the neuroimaging study (CDC/NIOSH U01 OH011314), the National Institute on Aging that supports research on characterization and treatment of Alzheimer’s disease (NIH/NIA P50 AG005138), and aging-relates work in this population (NIH/NIA R01 AG049953). We would also like to acknowledge ongoing funding to monitor World Trade Center responders as part of the WTC Health and Wellness Program (CDC 200- 2011-39361). This research was partially funded by the Swiss National Science Foundation (SNSF) Postdoc Mobility Fellowship (P500PB_206833). This work was supported in part through the computational and data resources and staff expertise provided by Scientific Computing and Data at the Icahn School of Medicine at Mount Sinai and supported by the Clinical and Translational Science Awards (CTSA) grant UL1TR004419 from the National Center for Advancing Translational Sciences. Research reported in this publication was also supported by the Office of Research Infrastructure of the National Institutes of Health under award number S10OD026880 and S10OD030463. The content is solely the responsibility of the authors and does not necessarily represent the official views of the National Institutes of Health. Additional support was provided by the Icahn School of Medicine Capital Campaign, BioMedical Engineering and Imaging Institute, and Department of Radiology, as well as by the Intramural Research Program of NINDS, NIH. This work was supported by the Office of the Assistant Secretary of Defense for Health Affairs through the Multiple Sclerosis Research Program under Award No. (HT9425-24-1-0857). Opinions, interpretations, conclusions, and recommendations are those of the author and are not necessarily endorsed by the Department of Defense.

## Supplementary Materials

**Table S1.** - Differences in brain volumes between WTC responders with and without PTSD. Only brain areas that resulted significantly correlated with PAD and predicted brain age were investigated.

| **Brain Area (Volume)** | **PTSD- (n=52)** | **PTSD+ (n=47)** | **p** |
| --- | --- | --- | --- |
| **X4th Ventricle** |  |  | 0.409 |
| mean ± sd | 0.00126 ± 0.0003 | 0.00132 ± 0.0003 | |
| **X3th Ventricle** |  |  | 0.012 |
| mean ± sd | 0.00061 ± 0.0002 | 0.00075 ± 0.0003 | |
| **Lateral Ventricle** |  |  | 0.674 |
| mean ± sd | 0.0074 ± 0.0032 | 0.0077 ± 0.0023 | |
| **Inferior Lateral Ventricle** |  |  | 0.347 |
| mean ± sd | 0.0004 ± 0.00014 | 0.00043 ± 0.0001 | |
| **Cerebrospinal fluid (CSF)** |  |  | 0.050 |
| mean ± sd | 0.2413 ± 0.0197 | 0.2489 ± 0.0182 | |
| **Thalamus** |  |  | 0.323 |
| mean ± sd | 0.00472 ± 0.0003 | 0.00465 ± 0.0003 | |

**Note:** Mean, standard deviation (sd) of brain volumes adjusted for total intracranial volume of each responder are reported. P-values quantifying differences between WTC-PTSD and non-PTSD were derived using Student *t*-tests. \**p*<0.005.

**Figure S1.**
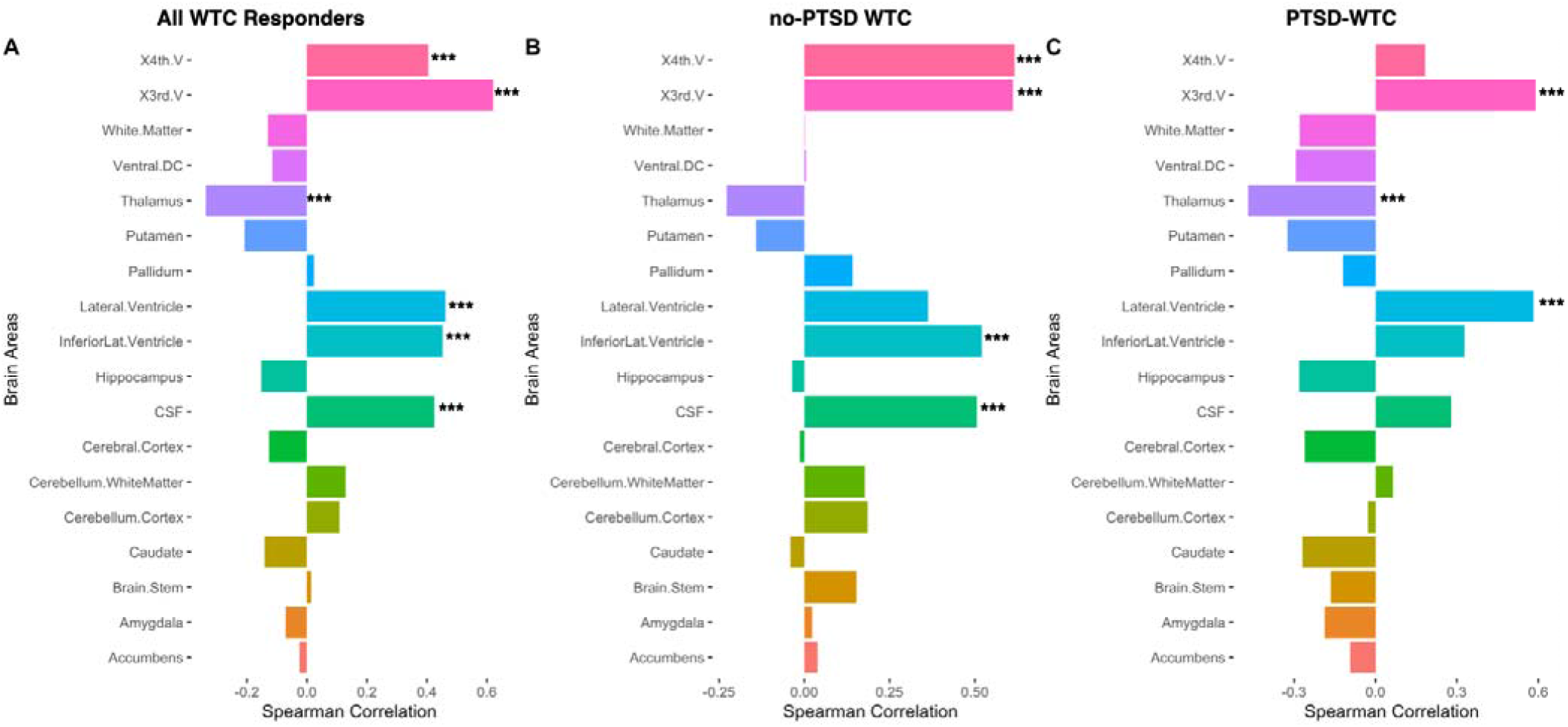
Predicted Brain age metrics are strongly correlated with specific brain areas. Spearman correlations between brain volumes (normalized for intracranial brain volume) and PAD for all 99 WTC responders (Panel A), non-PTSD (Panel B) and WTC-PTSD (Panel C). Brain parcellation was performed with SynthSeg (FreeSurfer)^58^. Significance level: * *p* < 0.05, ** *p* < 0.01, *** *p* < 0.001.

**Figure S2.**
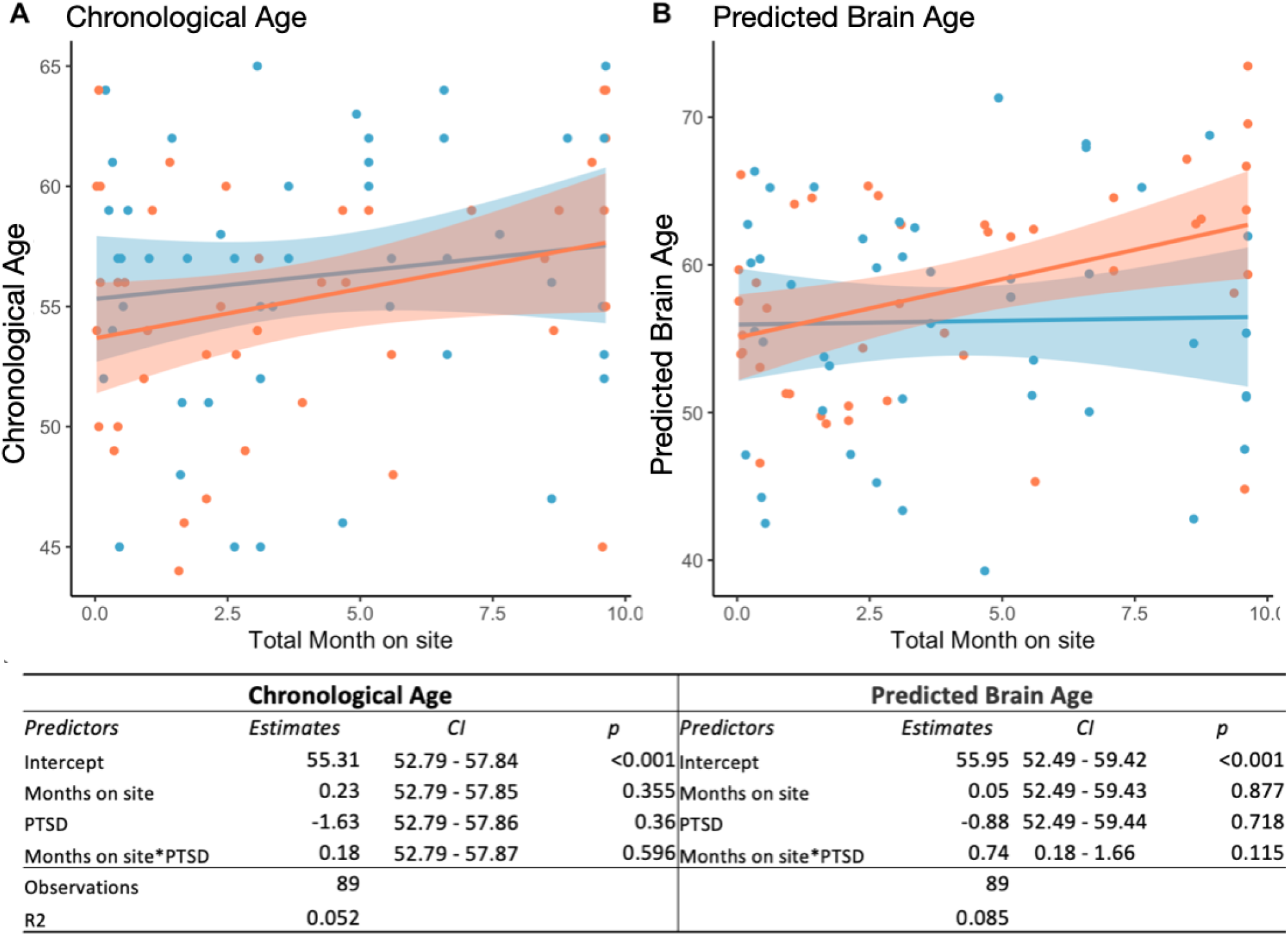
- WTC exposure duration, chronological age and predicted brain age. This graph plots the relationship (interaction) between WTC exposure duration in months (x-axis) and chronological age (Panel A) and predicted brain age (Panel B) (y-axis) stratified by WTC-PTSD (orange dots) and non-PTSD (blue dots).

